# Health outcome priorities of people with multiple long-term conditions before and during the COVID-19 pandemic: Survey data from the UK

**DOI:** 10.1101/2024.03.24.24304807

**Authors:** Harini Sathanapally, Yogini V Chudasama, Francesco Zaccardi, Alessandro Rizzi, Samuel Seidu, Kamlesh Khunti

## Abstract

**Background:** The outcome prioritisation tool (OPT) is a simple tool to ascertain the health outcome priorities of people with MLTC. Use of this tool in people aged under 65 years with MLTC has not previously been investigated. This study investigated the feasibility of using the OPT in people with MLTC aged 45 years or above, in a multi-ethnic primary-care setting, to describe the health outcome priorities of people with MLTC by age, clusters of long-term conditions and demographic factors, and to investigate any differences in prioritisation in light of the COVID-19 pandemic.

**Methods:** This was a multi-centre cross-sectional study using a questionnaire for online self-completion by people aged 45 years or above with MLTC in 19 primary care settings across the East Midlands, UK. Participants were asked to complete the OPT twice, first from their current perspective and second from their recollection of their priorities prior to COVID-19.

**Results:** The questionnaire was completed by 2,454 people with MLTC. The majority of participants agreed or strongly agreed that the OPT was easy to complete, relevant to their healthcare and will be useful in communicating priorities to their doctor. Summary scores for the whole cohort of participants showed Keeping Alive and Maintaining Independence receiving the highest scores. Statistically significant differences in prioritisation by age, clusters of long-term conditions and employment status were observed, with respondents aged over 65 most likely to prioritise Maintaining independence, and respondents aged under 65 most likely to prioritise Keeping alive. There were no differences before or after COVID-19, or by ethnicity.

**Conclusions:** The OPT is feasible and acceptable for use to elicit the health outcome priorities of people with MLTC across both middle-aged and older age groups and in a UK setting. Individual factors could influence the priorities of people with MLTC and must be considered by clinicians during consultations.

**HIGHLIGHTS:**

- Survey data from 2,454 patients with MLTC showed that keeping alive and maintaining independence were the top first-choice priorities from the health outcome priorities tool (OPT).
- The health outcomes priorities differed by socio-demographics and clusters of long-term conditions.
- There were no differences in health outcomes priorities before and during COVID-19.
- OPT is easy and acceptable to implement in a health care setting in a broad patient group.
- Translation of the OPT into different languages is recommended to address any potential language barrier for people with MLTC completing the OPT

## INTRODUCTION

Finding ways to improve the management of people with multiple long-term conditions (MLTC), also known as multimorbidity (1), is now a global priority for policy and healthcare research (2). The delivery of person-centred care, with the incorporation of patients’ priorities and preferences into decision-making, is key to the effective management of people with MLTC (3). The National Institute of Health and Care Excellence (NICE) highlights the inclusion of patient’s priorities, values and goals, in their quality standards for the management of people with MLTC. One previous systematic review showed there were major differences in the processes of prioritisation of people with MLTC and from the clinicians who were managing people with MLTC, thus highlighting the need to elicit the priorities of people with MLTC in each consultation, to avoid the risk of a misalignment of priorities (4). The review also found large variations in how priorities were ascertained, along with the tools used for this purpose, and we emphasised the need for a standardised tool to facilitate clinicians in eliciting the priorities of patients with MLTC (4).

The outcome prioritisation tool (OPT) previously developed by Fried et al, emerged as the most commonly used validated tool to date in our systematic review (4, 5). The OPT is a simple tool to facilitate clinicians in ascertaining the health outcome priorities of their patients across four domains, namely: maintaining independence, keeping alive, reducing pain, or reducing other symptoms (5). The feasibility of using the OPT to elicit the priorities of people with MLTC aged 65 years and over (5, 6), to facilitate medication review (7, 8), and complex treatment decision-making in older people with MLTC (9), has previously been investigated in clinical settings in the United States and in the Netherlands. However, this tool is yet to be applied to a population in the United Kingdom (UK), to younger people aged under 65 years or to an ethnically diverse population. Whilst a higher proportion of older adults suffer from MLTC (10), the prevalence of MLTC in younger adults is also high and continuing to rise (11). Ethnicity-based differences in the risk of diagnoses of MLTC have also been found, with people of Asian ethnicity at increased risk of suffering from MLTC (12). However there has been very little understanding of the health outcome priorities of people with MLTC aged under 65, and/or people with MLTC from ethnic minority backgrounds, and how their health outcome priorities can be elicited. Moreover, the COVID-19 pandemic raised further challenges for people with MLTC, as the presence of MLTC is an independent risk factor for an increased risk of severe COVID-19 and mortality (13,14).

Therefore, the aims of this study were to describe the health outcome priorities of multi-ethnic people with MLTC including those aged 65 years and under, investigate the association between patient’s current first-choice health priority and risk factors, compare the health outcome priorities of patients before the onset of the COVID-19 outbreak to their current health outcome priorities. A further aim was, to examine the feasibility of the validated OPT by patient-reported relevance, ease of use, and patient-perceived usefulness.

## METHODS

### Study design

This was a multi-centre, cross-sectional study using a questionnaire for self-completion by people aged 45 years and above and suffering from at least two long-term conditions in primary care across the East Midlands, UK. Data was collected from August 2020 to August 2022. NHS Ethical approval for this study was granted by the Riverside REC Committee (Reference:20/LO/0570). The survey required about 10 minutes for respondents to fill out and assessed participants’ demographic information, encompassing age, sex, ethnicity, self-reported long-term medical conditions, the quantity of regularly prescribed medications and self-reported status of NHS identification as being at very high risk or extremely vulnerable from Covid-19. The questionnaire was initially to be completed by participants in a paper format after face-to-face recruitment; however, due to the COVID-19 pandemic, the questionnaire was adapted to an online format.

Potential participants were identified by members of the direct care team in the participating primary care practices using an electronic search of practice records for people aged 45 or above and had two or more coded diagnoses of long-term conditions, including cardiovascular, respiratory, metabolic, mental health, neurological, musculoskeletal and chronic pain conditions. Eligible participants were sent a brief invitation message introducing the study, a link to the participant information sheet and the online questionnaire via text or e-mail by the administration teams. The responses were captured and stored in a secure database through the REDCap software (15). We did not collect any identifiable data from participants and therefore the completed questionnaires were fully anonymised from the outset.

### Health outcome prioritisation tool

Participants’ health outcome priorities were measured using the OPT (5), namely: maintaining independence, keeping alive, reducing pain, and reducing other symptoms. Participants were asked to prioritise and rank the four health outcomes using a scale of 0-100, where 0 is the least priority and 100 is the high priority. They were asked to complete the tool twice, first considering their current priorities and second considering their priorities before the COVID-19 pandemic. The health outcome priorities have previously shown outstanding reliability on prior testing but the VAS scores of 0-100 demonstrated substantial variation, therefore for the analysis we chose a priori to analyse rank orderings only, i.e., first-choice only (16). Due to the principle of trade-offs, if participants ranked two outcomes as the same i.e., 100, 100, or had any missing outcomes, these data were excluded from the analysis for the rankings to adhere to the OPT goals (17), hence this is the reduced cohort of the study population. After completing the OPT, participants were then asked to complete three Likert scale questions (ranging from strongly agree to strongly disagree) to measure the relevance, ease of use and patient-perceived usefulness of the OPT.

### Patient and public involvement engagement

We conducted a patient and public involvement (PPI) focus group with participants with MLTC aged 45 years or above and from a diverse range of ethnic backgrounds, to review the study documentation, including the participant information sheet and questionnaire. The purpose of this session was to gather input on the clarity and comprehensibility of these documents, as well as to gather any recommendations for enhancements. The feedback obtained from this session was used to amend the questionnaire to improve its readability and ease of understanding.

### Statistical analysis

The characteristics of the respondents and self-reported long-term conditions were presented as number (%), mean [standard deviation (SD)], or median [interquartile range (IQR)]. We described the health outcome priority scores in the whole cohort using the median [IQR]. In those who applied the trade-off principle (reduced cohort), we then examined the proportions of the rankings given as first-choice from the four health outcomes with further breakdown by the sociodemographic factors and clusters of conditions and the differences were tested using chi-squared statistics. The association between the respondent’s first-choice health priority and sociodemographic factors and clusters of long-term conditions was estimated using logistic regression models. The odds ratio (OR) and 95% confidence intervals (CI) were reported comparing each factor and cluster of condition by each of the four health priority outcomes as first-choice, where an OR <1 indicated a lower health priority, whereas OR >1 indicated a higher health priority. Unadjusted and adjusted models for age (continuous) and gender (female, male) were reported. To assess whether the health outcomes differed before COVID-19 and the current choice, we compared the continuous before and after scores for each outcome using the Wilcoxon signed ranked test. The feasibility analysis was carried out in the whole cohort which tested peoples’ ‘perceived usefulness’ of the OPT. The three Likert scale responses ranged from strongly agree, agree, neither agree nor disagree, disagree, or strongly disagree, this was compared using chi-squared statistics. All statistical analyses were performed in Stata/IC 17.0; results were reported with 95% CI and statistical significance was defined at P<0.05. The statistical code is available at GitHub *yc244*.

## RESULTS

### Survey respondents

Overall 2,454 people with MLTC completed the survey from 19 primary care practices across the East Midlands, UK, **Figure S1**. The mean [SD] age was 63.6 [10.1] years, 57.1% were aged 45-65 years, and 42% were aged over 65 years; 58% were female, 92% were of White ethnicity, 36% were working, 8% were unemployed and 47% were retired, **Table 1**. A further breakdown of these categories is provided in **Table S1**. The median [IQR] number of self-reported long-term conditions was 3 [IQR 2 - 4], with the most common conditions being hypertension (48%), arthritis (36%), chronic pain (29%), depression (27%), anxiety (27%), and diabetes (26%), **Table S2**. When examining the clusters of long-term conditions, the largest was for cardiometabolic conditions (65%), musculoskeletal or chronic pain (52%), mental health (36%), and respiratory diseases (33%), **Table 1**. Regular medications were taken by 94% of the cohort, with the median [IQR] number of medications being 5 [3-7]. Almost 70% were at high risk of COVID-19, **Table 1**.

**Table 1.**
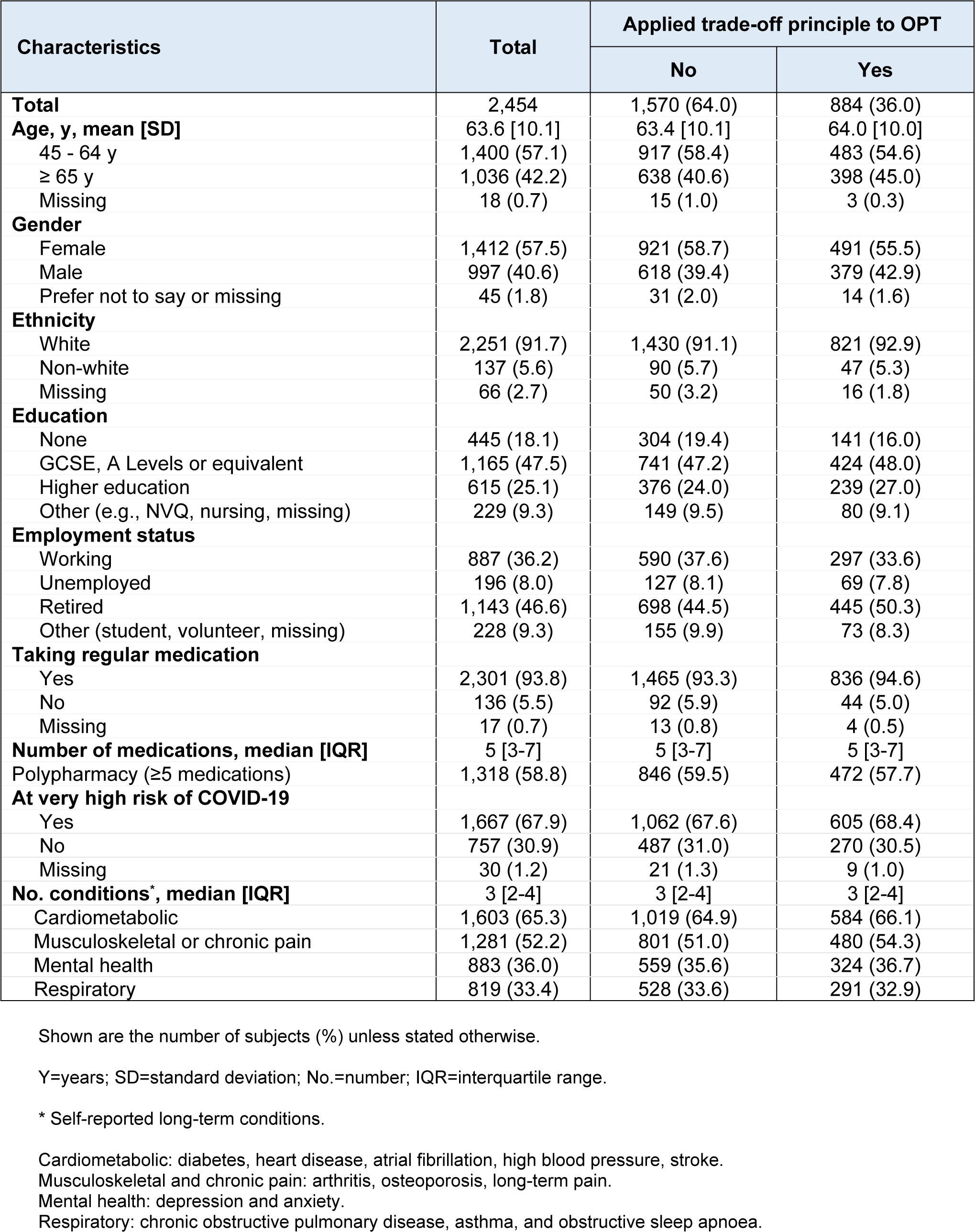
Characteristics of all survey respondents.

| Characteristics | Total | Applied trade-off principle to OPT |  |
| --- | --- | --- | --- |
|  |  | No | Yes |
| **Total** | 2,454 | 1,570 (64.0) | 884 (36.0) |
| **Age, y, mean [SD]** | 63.6 [10.1] | 63.4 [10.1] | 64.0 [10.0] |
| 45 - 64 y | 1,400 (57.1) | 917 (58.4) | 483 (54.6) |
| ≥ 65 y | 1,036 (42.2) | 638 (40.6) | 398 (45.0) |
| Missing | 18 (0.7) | 15 (1.0) | 3 (0.3) |
| **Gender** |  |  |  |
| Female | 1,412 (57.5) | 921 (58.7) | 491 (55.5) |
| Male | 997 (40.6) | 618 (39.4) | 379 (42.9) |
| Prefer not to say or missing | 45 (1.8) | 31 (2.0) | 14 (1.6) |
| **Ethnicity** |  |  |  |
| White | 2,251 (91.7) | 1,430 (91.1) | 821 (92.9) |
| Non-white | 137 (5.6) | 90 (5.7) | 47 (5.3) |
| Missing | 66 (2.7) | 50 (3.2) | 16 (1.8) |
| **Education** |  |  |  |
| None | 445 (18.1) | 304 (19.4) | 141 (16.0) |
| GCSE, A Levels or equivalent | 1,165 (47.5) | 741 (47.2) | 424 (48.0) |
| Higher education | 615 (25.1) | 376 (24.0) | 239 (27.0) |
| Other (e.g., NVQ, nursing, missing) | 229 (9.3) | 149 (9.5) | 80 (9.1) |
| **Employment status** |  |  |  |
| Working | 887 (36.2) | 590 (37.6) | 297 (33.6) |
| Unemployed | 196 (8.0) | 127 (8.1) | 69 (7.8) |
| Retired | 1,143 (46.6) | 698 (44.5) | 445 (50.3) |
| Other (student, volunteer, missing) | 228 (9.3) | 155 (9.9) | 73 (8.3) |
| **Taking regular medication** |  |  |  |
| Yes | 2,301 (93.8) | 1,465 (93.3) | 836 (94.6) |
| No | 136 (5.5) | 92 (5.9) | 44 (5.0) |
| Missing | 17 (0.7) | 13 (0.8) | 4 (0.5) |
| **Number of medications, median [IQR]** | 5 [3-7] | 5 [3-7] | 5 [3-7] |
| Polypharmacy (≥5 medications) | 1,318 (58.8) | 846 (59.5) | 472 (57.7) |
| **At very high risk of COVID-19** |  |  |  |
| Yes | 1,667 (67.9) | 1,062 (67.6) | 605 (68.4) |
| No | 757 (30.9) | 487 (31.0) | 270 (30.5) |
| Missing | 30 (1.2) | 21 (1.3) | 9 (1.0) |
| **No. conditions\*, median [IQR]** | 3 [2-4] | 3 [2-4] | 3 [2-4] |
| Cardiometabolic | 1,603 (65.3) | 1,019 (64.9) | 584 (66.1) |
| Musculoskeletal or chronic pain | 1,281 (52.2) | 801 (51.0) | 480 (54.3) |
| Mental health | 883 (36.0) | 559 (35.6) | 324 (36.7) |
| Respiratory | 819 (33.4) | 528 (33.6) | 291 (32.9) |

Shown are the number of subjects (%) unless stated otherwise.
Y=years; SD=standard deviation; No.=number; IQR=interquartile range.
\* Self-reported long-term conditions.
Cardiometabolic: diabetes, heart disease, atrial fibrillation, high blood pressure, stroke.
Musculoskeletal and chronic pain: arthritis, osteoporosis, long-term pain.
Mental health: depression and anxiety.
Respiratory: chronic obstructive pulmonary disease, asthma, and obstructive sleep apnoea.

Overall, 64% (n=1,570) of respondents were not able to apply the trade-off principle to all the health outcomes as they either assigned the same score to more than one outcome or had a missing outcome. 36% (n=884) of participants were able to apply the trade-off principle and gave a hierarchy of scores to all the health outcomes, **Figure S1**. The characteristics of those who did and did not apply the trade-off principle were similar, **Table 1**.

### Summary of health outcome priority scores

In the whole cohort (N=2,454), the health outcome of keeping alive was the highest priority with the median value of 98 [IQR 88 - 100], followed by maintaining independence (97 [85 - 100]), reducing pain (92 [77 - 99]), and reducing other symptoms (87 [74 - 99]), **Figure 1**.

**Figure 1.**
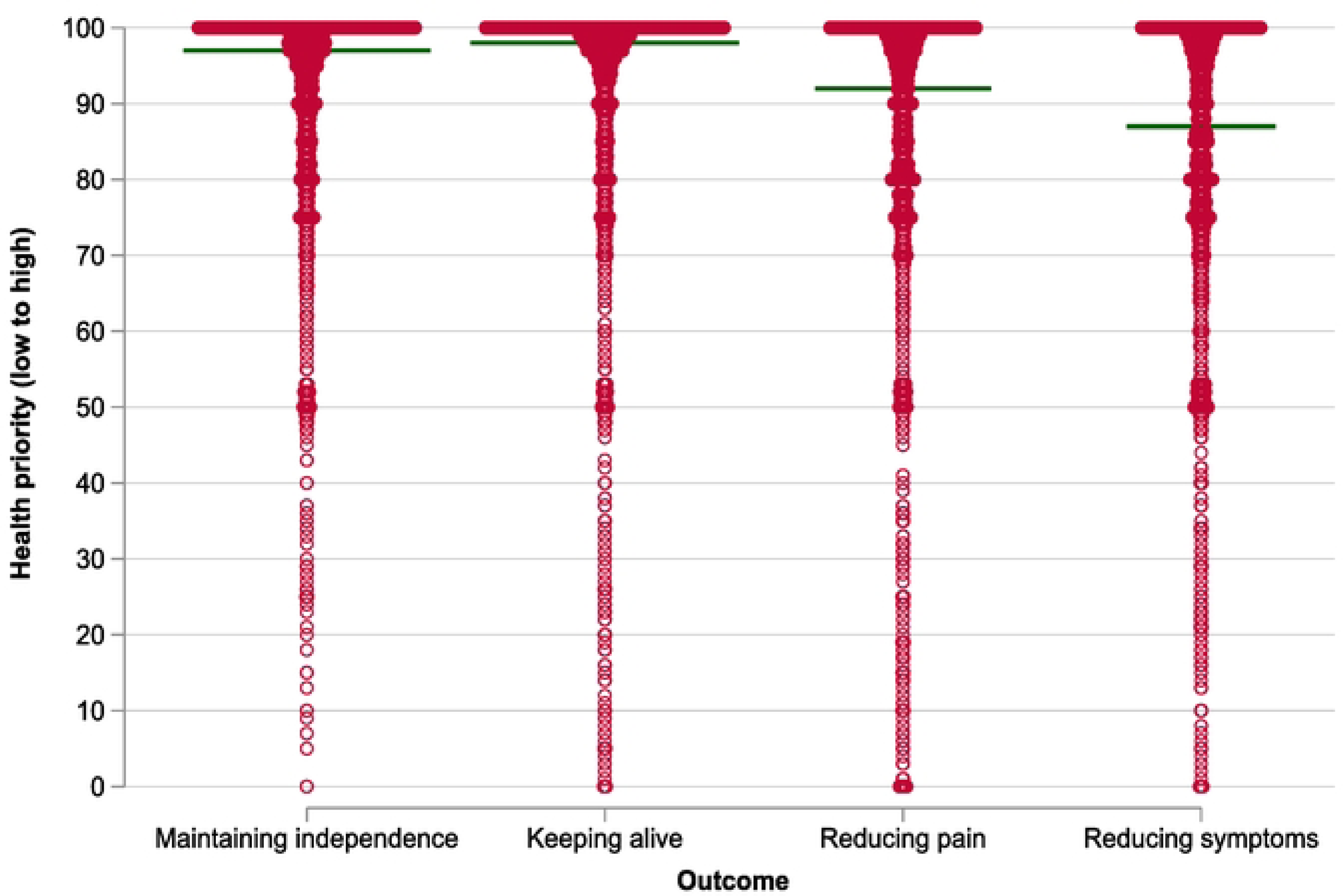
Health outcome priority score in the whole cohort. Reference line represents the median value of health prioritisation on the scale of 0 to 100. Median [interquartile range]: maintaining independence 97 [85 - 100], keeping alive 98 [88 - 100], reducing pain 92 [77 - 99], reducing other symptoms 87 [74 - 99]. Missing outcome: maintaining independence 140, keeping alive 164, reducing pain 221, reducing other symptoms 269.

### Analysis of first-choice health outcome priority

#### Current first-choice health outcome priority

In the cohort of participants who did apply the trade-off principle, (n=884), keeping alive was the outcome top-ranked as first-choice (44%), next was maintaining independence (33%), then reducing pain (14%), and reducing other symptoms (10%), **Table S3**. When comparing by the respondent’s sociodemographic factors and clusters of conditions, we found statistically significant differences between most groups, **Table S3**. This was further illustrated by the significant associations **(Table S4, Table S5, Figure 2).** Maintaining independence was highly prioritised for those aged 65 years or more (adjusted OR 1.55 (95% CI 1.16, 2.06)) compared to aged 45-65 years; with higher education (2.27 (1.60, 4.14)), or secondary/further education (1.73 (1.11, 2.71)), compared with no education; and those at high risk of COVID-19 (1.60 (1.15, 2.21)). Male respondents were 37% more likely to prioritise keeping alive compared to females (1.37 (1.04, 1.80)).

**Figure 2.**
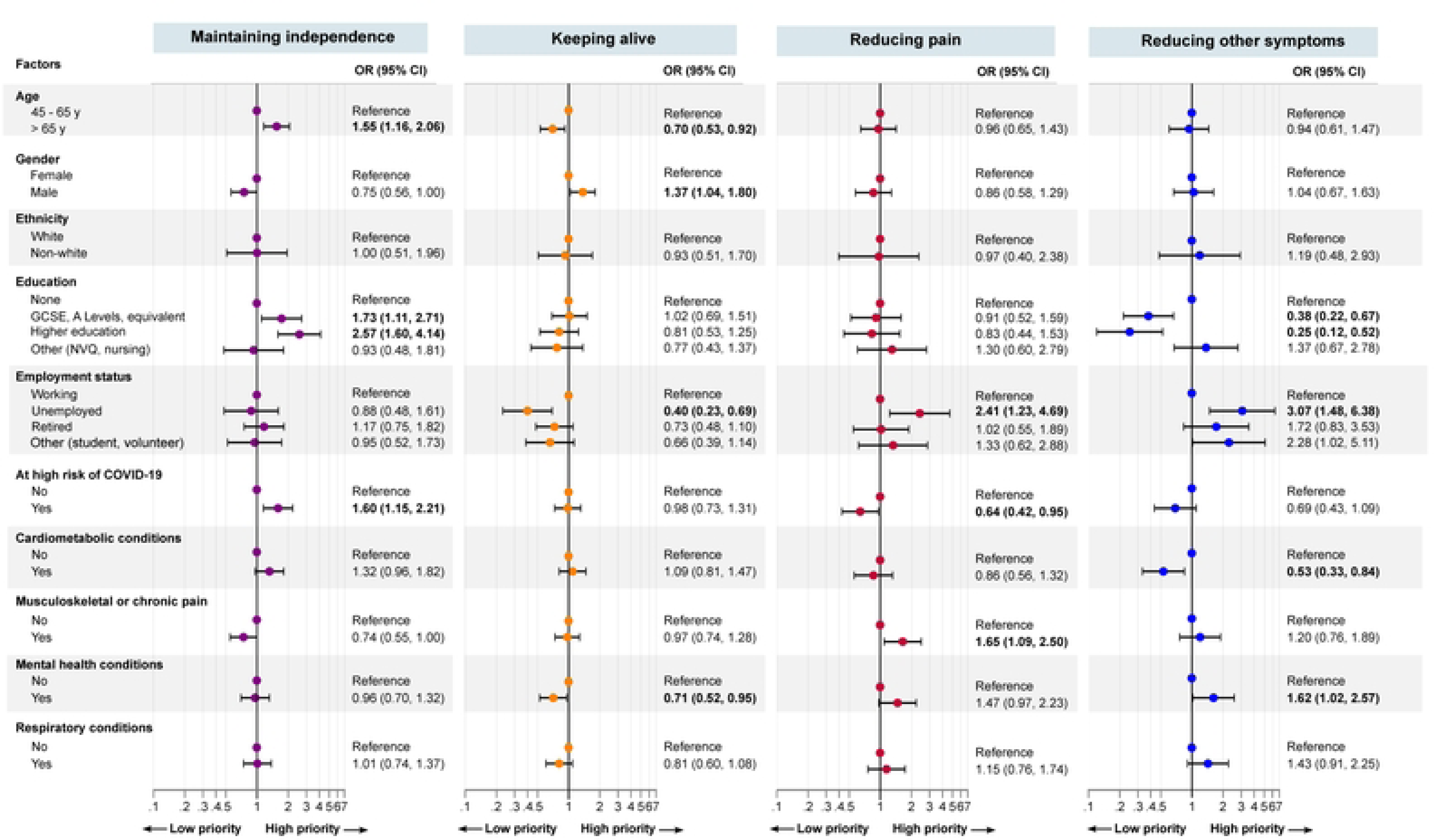
Association between patient’s first-choice health priority and sociodemographic factors and clusters of long-term conditions. Models are adjusted by age (continuous) and gender (female or male). CI=confidence interval. Odds ratio <1 indicates low health priority, whereas odds ratio >1 indicate high health priority. Bold indicates statistical significance, P<0.05.

Respondents who were unemployed prioritised reducing other symptoms three times more than those working (3.07 (1.48, 6.38), and reducing pain two times more [(2.41 (1.23, 4.69), and least prioritised keeping alive (0.40 (0.23, 0.69)). For those with musculoskeletal or chronic pain, their first-choice priority was reducing pain (1.65 (1.09, 2.50), and their least priority was maintaining independence (0.74, (0.55, 1.00)). While respondents who reported mental health conditions prioritised reducing other symptoms (1.62 (1.02, 2.57)), and least prioritised keeping alive (0.71 (0.52, 0.95)). Comparison by respondents’ ethnicity did not reveal any statistically significant differences in prioritisation **Table S5, Figure 2**.

#### Comparing the first-choice health outcome priorities before COVID-19 to current

From a “before COVID-19” perspective the outcome most frequently ranked as the first-choice was keeping alive (38%), next maintaining independence (34%), then reducing pain (18%), and reducing other symptoms (10%), **Table S6**. There were no significant differences in health outcome priority before COVID-19 and current using the continuous scores, further evidenced in **Figure S2** and **Figure S3**.

### Feasibility of the health outcome prioritisation tool

In the whole cohort, the majority of respondents agreed or strongly agreed that the OPT was easy to complete (68%), was relevant to their healthcare (57%), and useful in communicating what their priorities are to their doctor (60%), **Table 2**. Similar feasibility results were found in those who did and did not apply the trade-off principle, **Table 2**. In those who applied the trade-off principle (n=884), the tool was particularly favoured by respondents with primary or secondary school education, at high risk of COVID-19, and with cardiometabolic conditions, musculoskeletal conditions or chronic pain (**Table S7, Table S8, Table S9**).

**Table 2.** Feasibility questions of patient’s perceived usefulness of the health outcome prioritisation tool.

| Feasibility question | Total<br>N=2,454 | Applied trade-off principle to HOPT |  |
| --- | --- | --- | --- |
|  |  | No<br>N=1,570 | Yes<br>N=884 |
| **1) The health outcome prioritisation tool was easy to complete** |  |  |  |
| Strongly agree | 563 (22.9) | 400 (25.5) | 163 (18.4) |
| Agree | 1,108 (45.2) | 697 (44.4) | 411 (46.5) |
| Neither agree nor disagree | 489 (19.9) | 305 (19.4) | 184 (20.8) |
| Disagree | 147 (6.0) | 71 (4.5) | 76 (8.6) |
| Strongly disagree | 133 (5.4) | 83 (5.3) | 50 (5.7) |
| Missing | 14 (0.6) | 14 (0.9) | 0 (0.0) |
| **2) The health outcome prioritisation was relevant to my healthcare** |  |  |  |
| Strongly agree | 308 (12.6) | 232 (14.8) | 76 (8.6) |
| Agree | 1,089 (44.4) | 675 (43.0) | 414 (46.9) |
| Neither agree nor disagree | 772 (31.5) | 487 (31.0) | 285 (32.3) |
| Disagree | 183 (7.5) | 111 (7.1) | 72 (8.2) |
| Strongly disagree | 88 (3.6) | 53 (3.4) | 35 (4.0) |
| Missing | 14 (0.6) | 12 (0.8) | 2 (0.2) |
| **3) The health outcome prioritisation tool will be useful in communicating what my priorities are to my doctor** |  |  |  |
| Strongly agree | 378 (15.4) | 292 (18.6) | 86 (9.7) |
| Agree | 1,092 (44.5) | 652 (41.5) | 440 (49.8) |
| Neither agree nor disagree | 639 (26.0) | 401 (25.5) | 238 (26.9) |
| Disagree | 217 (8.8) | 140 (8.9) | 77 (8.7) |
| Strongly disagree | 118 (4.8) | 75 (4.8) | 43 (4.9) |
| Missing | 10 (0.4) | 10 (0.6) | 0 (0.0) |

## DISCUSSION

Our study describes the health outcome priorities of a large cohort of participants and compares the health outcome priorities by age, clusters of long-term conditions, ethnicity and demographic factors. To our knowledge, this is the largest study to investigate the use of the OPT in people with MLTC, the first study to investigate the use of the OPT in the UK, in a multi-ethnic population, and in people with MLTC aged under 65. This is also the first study to compare the health outcome priorities of middle-aged people with MLTC (aged 45 to 65) and older people with MLTC (aged over 65), and whether the health outcome priorities of people with MLTC have changed in light of the COVID-19 pandemic. This study has made a novel contribution to understanding the health outcome priorities of people with MLTC and the factors that may influence them. This is a step towards facilitating the development of priorities-based models of care for people with MLTC.

With a mean participant age of 64 years, the cohort in our study can be considered to be representative of middle-aged people (18) with MLTC. The majority of participants agreed or strongly agreed that the OPT was easy to complete, relevant to their healthcare and will be useful in communicating priorities to their doctor. Hence, the results show that the OPT is relevant, useful, and easy to complete for middle-aged people with MLTC.

Overall, we found *keeping alive* was most frequently ranked as the first-choice health outcome priority, with *maintaining independence* being second most likely to be ranked as the top priority. Previous studies using the OPT in people with MLTC with sample sizes ranging from 59 to 357, have found that *maintaining independence* was most frequently ranked as the top priority (5, 8, 19) (7). This difference from our overall findings could be due to the fact that previous studies using the OPT have all focused on older people with MLTC, whereas we included middle-aged people with MLTC in our cohort. Indeed, comparison of participants’ ranking by age, revealed that participants aged over 65 were most likely to rank *maintaining Independence* as their top priority, and were more likely to do so than participants age under 65 years, which is in keeping with the findings of previous literature (5, 8, 19).

This age-based difference in the choice of first-choice health outcome priority in our findings further demonstrates that age may influence how people with MLTC prioritise their health outcomes. A previous systematic review also demonstrated age-based differences in how clinicians arrived at health outcome and treatment priorities for people with MLTC, as some clinicians reported that prioritising reduced the risk of mortality more highly as they felt they could be “more aggressive” in their clinical management, in people with MLTC aged under 65 (4).

We also observed statistically significant differences in participants’ priorities based on types of long-term conditions. Participants with musculoskeletal conditions or chronic pain were most likely to choose *reducing pain* as their first-choice health outcome priority. Previous literature has demonstrated that the empirical impact of illnesses including their symptom burden (4), and disease burden (20), were factors which influenced how people with MLTC chose priorities related to their health, and could provide the likely explanation for this finding, as the symptom of pain has the potential to form a significant part of the symptom burden experienced by people with musculoskeletal conditions (21), and/or chronic pain. Participants with mental health conditions were most likely to choose r*educing other symptoms* as their top priority, and *keeping alive* the least, which suggests that for these participants, their health outcome priorities could be related to the symptom burden of their illness but these were not available as a choice to be captured on the OPT.

We also found that socio-economic factors such as education and employment status were associated with statistically significant differences in prioritisation. Adjusting for age and gender, our results indicated that participants who were unemployed were three times more likely to prioritise *reducing other symptoms*, and twice more likely to prioritise *reducing pain*. Prolonged unemployment has previously been found to be associated with musculoskeletal pain (22). A previous study in the context of multiple sclerosis found that the presence of pain was associated with a reduced rate of employment (23), and another study investigating the impact of pain on employment amongst cancer survivors found that pain was associated with an increased risk of adverse employment outcomes (24). Our findings demonstrate that socio-economic factors can have an impact on the health outcome priorities of people with MLTC, and highlight the importance of taking a holistic person-centred approach that considers the impact of the socio-economic background of people with MLTC on an individual basis when eliciting their health outcome priorities in consultations.

We did not observe any statistically significant changes in participants’ reporting of their health outcome prioritisation from before the COVID-19 pandemic and currently. However, these results are subject to recall bias, as participants were asked to retrospectively report their health outcome priorities from before the onset of COVID-19.

The OPT has previously been used in a face-to-face format, where participants were helped by facilitators to indicate their order of prioritisation (5). However, in this study, the OPT was disseminated in an online questionnaire format in light of the COVID-19 pandemic, and whilst participants were advised in the participant information sheet to contact their practice or the study team if they had any questions regarding the study, direct facilitation to help patients complete the OPT was not feasible. We found that a significant number of participants (64%) gave the same scores to multiple domains or missed outcomes, raising the possibility that the “trade-off” principle with using the OPT may not have been clearly understood by participants on these occasions. The online format of the questionnaire, without the input of a facilitator or clinician, could have been a contributing factor to this.

The fact that participants were being asked to consider their priorities outside of a situation where there was an actual decision to be made rather than a hypothetical one, could also have been a contributing factor to this. Indeed, a previous study investigating the use of the OPT for medication review for older people (aged 69 years or above) with MLTC found that participants reported engaging with prioritisation to be “difficult when there was no specific need to make a decision” (7).

We suggest that the OPT could be sent remotely to people with MLTC’s to complete ahead of their consultations, with their responses being reviewed again with their clinicians during their consultations to clarify that the trade-off principle has been applied, and the responses being revisited at any point where there is a management or treatment decision to be made. This approach would provide people with MLTC’s with time and opportunity to consider their health outcome priorities ahead of their appointments and discuss these with their clinicians during their appointments, which could promote the efficacy and efficiency of prioritised-based consulting and are, in primary care settings.

## STRENGTHS AND LIMITATIONS

To our knowledge, this was the first large study to implement the OPT in a UK setting, and the first to implement the OPT in participants from a multi-ethnic population aged under 65 years. With participation from multiple primary care practices across the East-midlands, and with a participant cohort representative of middle-aged and older people with MLTC, our study findings have demonstrated that people with MLTC found the OPT easy to use, relevant to their healthcare and perceive it to be useful in communicating their health outcome priorities to their doctor. Therefore, we found that this simple tool is acceptable to people with MLTC aged 45 years or above, and can feasibly be used in a primary care setting in future interventions to facilitate priorities-based care for people with MLTC in a primary care setting.

However, there are noteworthy limitations. As this was a cross-sectional study conducted after the onset of the COVID-19 pandemic, participants’ responses regarding their health outcome priorities prior to the COVID-19 pandemic were retrospective and hence subject to recall bias. We also found that a significant number of participants assigned the same scores to multiple health outcomes, suggesting that the “trade-off” principle was not always understood by participants, which could have been due to the virtual format of the questionnaire and the absence of a facilitator. The need to assign a score to each health outcome as part of the tool could also potentially lead to confusion regarding the trade-off principle.

Despite the study being set in a multi-ethnic setting with recruitment of primary care practices with high levels of ethnic diversity amongst their practice populations, we noted that there was a relatively low uptake of participation in our study from participants from ethnic minority backgrounds, with 92% of participants being of White ethnicity, and lower proportions of ethnic diversity amongst participants compared to National and Regional demographic data (25). A possible explanation for this could be due to a language barrier, as the invitation to participate in the study, and the study questionnaire itself were not available in any other languages besides English at this stage.

## RECOMMENDATIONS FOR THE FUTURE

To reduce the risk of confusion in assigning scores to different health outcomes and facilitate participants in applying the trade-off principle to reflect a hierarchy of health outcomes in order of importance to them on an individual basis, we propose a re-design of the OPT in which participants are asked to place each health outcome into four coloured boxes in order of their individual importance to them (**Figure 3**). We propose that this redesign would clearly capture the rank assigned to each health outcome by the participant and mitigate the potential for confusion through the removal of a need for participants to assign a score. We also recommend translating the OPT into different languages to address any potential language barrier for people with MLTC completing the OPT. Additionally, we suggest that the OPT could be used remotely ahead of consultations with people with MLTC, with review of responses during the consultation to clarify that the trade-off principle has been applied, and incorporation into the consultation to facilitate priorities-based and person-centred management.

**Figure 3.**
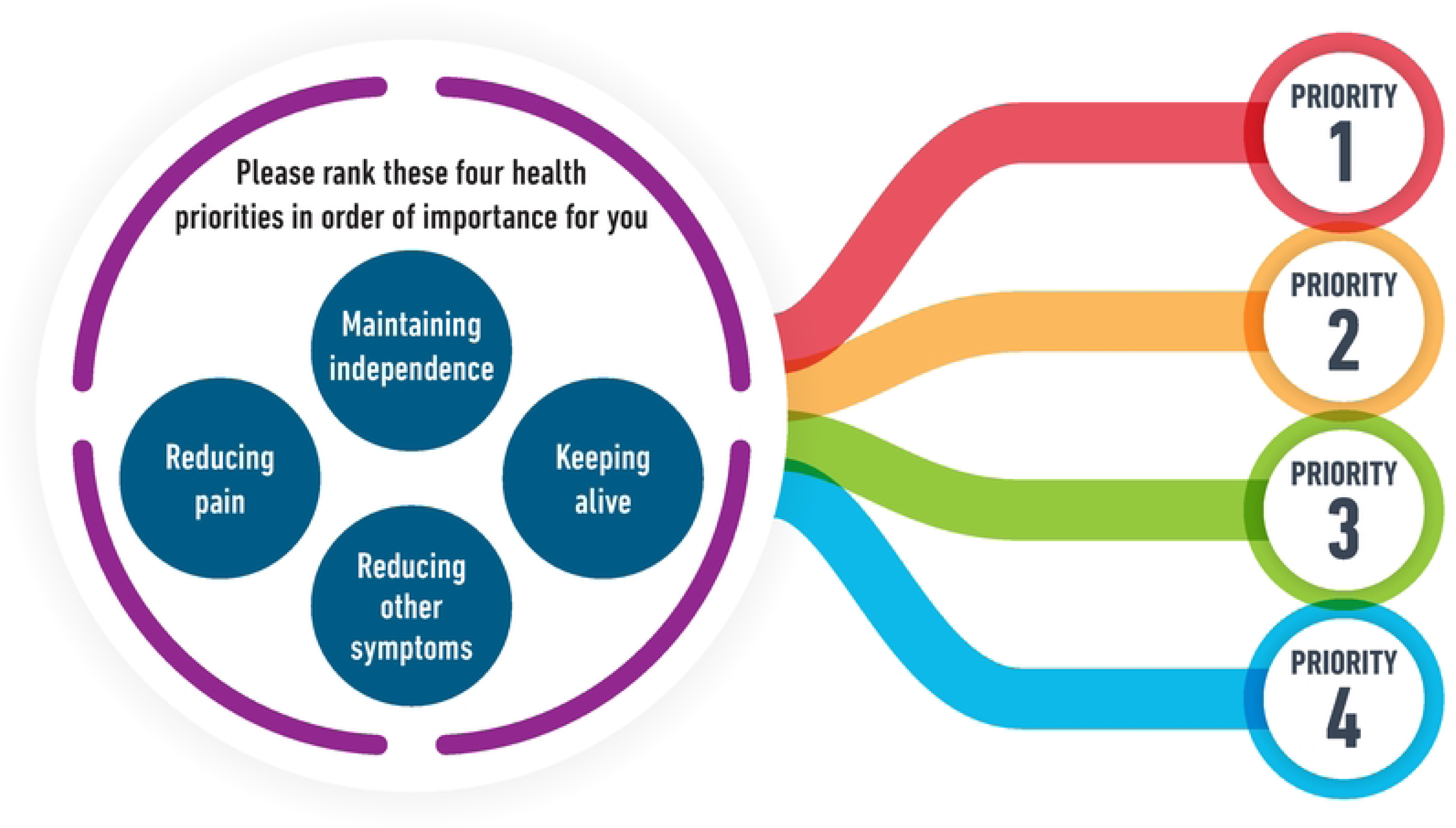
Health outcome prioritisation tool simplified for clinical use. Health outcome prioritisation tool developed by Fried et al (4).

## CONCLUSION

As a simple tool that encourages people with MLTC to consider and record what is important to them in terms of their health outcomes, we found the OPT is simple and acceptable by people with MLTC and has the potential to be a valuable means to introduce and incorporate individual priorities into their clinical consultations and facilitate person-centred, priorities-based care planning. We also found that the health outcome priorities of people with MLTC can be influenced by a number of factors individual to each patient such as age, types of long-term conditions, and employment status. We therefore recommend that in their consultations with people with MLTC, clinicians should take a holistic approach that takes the potential impact of all of these factors into account, in the planning and provision of priorities-based care for people with MLTC.

## DECLARATIONS

## Data Availability

All relevant data are within the manuscript and its Supporting Information files.

## Acknowledgements

We would like to thank the East Midlands CRN for their support with this study. KK is supported by the National Institute for Health Research (NIHR) Applied Research Collaboration East Midlands (ARC EM), NIHR Global Research Centre for Multiple Long-Term Conditions, MLTC Cross NIHR Collaboration (CNC) and the NIHR Leicester Biomedical Research Centre (BRC). SS, YC, FZ and HS are supported by NIHR ARC EM.

## Ethical approval

NHS Ethical approval for this study was granted by the Riverside REC Committee (Reference:20/LO/0570).

## Author Contribution

Conceptualization: HS, KK, SS Study design: HS, KK, SS Data preparation: HS, YC Data analysis: HS, YC, FZ, AR First draft: HS, YC, KK, SS Study critical revision and manuscript draft: All authors.

All authors provided final approval of the version to publish.

HS had full access to all the data in the study and had final responsibility for the decision to submit it for publication.

## ABBREVIATIONS

CI: Confidence Interval
CRF: Case Report Form
CT: Clinical Trials
EC: Ethics Committee (see REC)
GP: General Practice
OPT: Outcome prioritisation tool
ICF: Informed Consent Form
MLTC: Multiple long-term conditions
NHS: National Health Service
NRES: National Research Ethics Service
PPI: Patient and Public Involvement
R&D: NHS Trust R&D Department
REC: Research Ethics Committee
SD: Standard Deviation
UK: United Kingdom

## References

1. Khunti K, Sathanapally H, Mountain P. Multiple long term conditions, multimorbidity, and co-morbidities: we should reconsider the terminology we use. BMJ. 2023;383.

2. Academy of Medical Sciences. Multimorbidity: a priority for global health research 2018. 2018.

3. Muth C, van den Akker M, Blom JW, Mallen CD, Rochon J, Schellevis FG, et al. The Ariadne principles: how to handle multimorbidity in primary care consultations. BMC medicine. 2014;12(1):1–11.

4. Sathanapally H, Sidhu M, Fahami R, Gillies C, Kadam U, Davies MJ, et al. Priorities of patients with multimorbidity and of clinicians regarding treatment and health outcomes: a systematic mixed studies review. BMJ open. 2020;10(2):e033445.

5. Fried TR, Tinetti M, Agostini J, Iannone L, Towle V. Health outcome prioritization to elicit preferences of older persons with multiple health conditions. Patient Educ Couns. 2011;83(2):278–82.

6. Fried TR, Tinetti ME, Iannone L, O’Leary JR, Towle V, Van Ness PH. Health outcome prioritization as a tool for decision making among older persons with multiple chronic conditions. Arch Intern Med. 2011;171(20):1856–8.

7. van Summeren JJ, Haaijer-Ruskamp FM, Schuling J. Eliciting preferences of multimorbid elderly adults in family practice using an outcome prioritization tool. J Am Geriatr Soc. 2016;64(11):e143–8.

8. van Summeren JJ, Schuling J, Haaijer-Ruskamp FM, Denig P. Outcome prioritisation tool for medication review in older patients with multimorbidity: a pilot study in general practice. British Journal of General Practice. 2017;67(660):e501–6.

9. Festen S, van Twisk YZ, van Munster BC, de Graeff P. ‘What matters to you?’Health outcome prioritisation in treatment decision-making for older patients. Age Ageing. 2021;50(6):2264–9.

10. Barnett K, Mercer SW, Norbury M, Watt G, Wyke S, Guthrie B. Epidemiology of multimorbidity and implications for health care, research, and medical education: a cross-sectional study. The Lancet. 2012;380(9836):37-43.

11. Malecki SL, Van Mil S, Graffi J, Breetvelt E, Corral M, Boot E, et al. A genetic model for multimorbidity in young adults. Genetics in Medicine. 2020;22(1):132–41.

12. Valabhji J, Barron E, Pratt A, Hafezparast N, Dunbar-Rees R, Turner EB, et al. Prevalence of multiple long-term conditions (multimorbidity) in England: a whole population study of over 60 million people. J R Soc Med. 2023 Oct 31:1410768231206033.

13. Agrawal U, Azcoaga-Lorenzo A, Fagbamigbe AF, Vasileiou E, Henery P, Simpson CR, et al. Association between multimorbidity and mortality in a cohort of patients admitted to hospital with COVID-19 in Scotland. J R Soc Med. 2022;115(1):22–30.

14. Russell CD, Lone NI, Baillie JK. Comorbidities, multimorbidity and COVID-19. Nat Med. 2023;29(2):334–43.

15. Harris PA, Taylor R, Minor BL, Elliott V, Fernandez M, O’Neal L, et al. The REDCap consortium: Building an international community of software platform partners. J Biomed Inform. 2019;95:103208.

16. Ramer SJ, McCall NN, Robinson-Cohen C, Siew ED, Salat H, Bian A, et al. Health outcome priorities of older adults with advanced CKD and concordance with their nephrology providers’ perceptions. Journal of the American Society of Nephrology: JASN. 2018;29(12):2870.

17. Stegmann ME, Festen S, Brandenbarg D, Schuling J, van Leeuwen B, de Graeff P, et al. Using the Outcome Prioritization Tool (OPT) to assess the preferences of older patients in clinical decision-making: a review. Maturitas. 2019;128:49–52.

18. Medley ML. Life satisfaction across four stages of adult life. The International Journal of Aging and Human Development. 1980;11(3):193–209.

19. Fried TR, Tinetti ME, Iannone L, O’Leary JR, Towle V, Van Ness PH. Health outcome prioritization as a tool for decision making among older persons with multiple chronic conditions. Arch Intern Med. 2011;171(20):1856–8.

20. Cai M, Cui M, Nong Y, Qin J, Mo S. Health Priorities in Chronic Obstructive Pulmonary Disease Patients with Multimorbidity: A Qualitative Study. Patient preference and adherence. 2022:2521–31.

21. Picavet H, Schouten J. Musculoskeletal pain in the Netherlands: prevalences, consequences and risk groups, the DMC3-study. Pain. 2003;102(1-2):167–78.

22. Doku DT, Acacio-Claro PJ, Koivusilta L, Rimpelä A. Health and socioeconomic circumstances over three generations as predictors of youth unemployment trajectories. Eur J Public Health. 2019;29(3):517–23.

23. Shahrbanian S, Auais M, Duquette P, Anderson K, Mayo NE. Does pain in individuals with multiple sclerosis affect employment? A systematic review and meta-analysis. Pain research and management. 2013;18:e94–e100.

24. Halpern MT, de Moor JS, Yabroff KR. Impact of pain on employment and financial outcomes among cancer survivors. Journal of Clinical Oncology. 2022;40(1):24.

25. Office for National Statistics, National identity, England and Wales: Census 2021 [Internet]. [cited 13/12/22]. Available from: https://www.ons.gov.uk/peoplepopulationandcommunity/culturalidentity/ethnicity/bulletins/nationalidentityenglandandwales/census2021.

